# Diagnostic performance of the combined nasal and throat swab in patients admitted to hospital with suspected COVID-19

**DOI:** 10.1101/2020.10.03.20206243

**Authors:** Kuan Ken Lee, Dimitrios Doudesis, Daniella A. Ross, Anda Bularga, Claire L. MacKintosh, Oliver Koch, Ingolfur Johannessen, Kate Templeton, Sara Jenks, Andrew R. Chapman, Anoop S.V. Shah, Atul Anand, Meghan R. Perry, Nicholas L. Mills, on behalf of the DataLoch COVID-19 Collaboration

## Abstract

**Background:** Accurate diagnosis in patients with suspected coronavirus disease 2019 (COVID-19) is essential to guide treatment and limit spread of the virus. The combined nasal and throat swab is used widely, but its diagnostic performance is uncertain.

**Methods:** In a prospective, multi-centre, cohort study conducted in secondary and tertiary care hospitals in Scotland, we evaluated the combined nasal and throat swab with reverse transcriptase-polymerase chain reaction (RT-PCR) for severe acute respiratory syndrome coronavirus-2 (SARS-CoV-2) in consecutive patients admitted to hospital with suspected COVID-19. Diagnostic performance of the index and serial tests was evaluated for a primary outcome of confirmed or probable COVID-19, and a secondary outcome of confirmed COVID-19 on serial testing. The diagnosis was adjudicated by a panel, who recorded clinical, laboratory and radiological features blinded to the test results.

**Results:** We enrolled 1,369 consecutive patients (68 [53-80] years, 47% women) who underwent a total of 3,822 tests (median 2 [1-3] tests per patient). The primary outcome occurred in 36% (496/1,369), of whom 65% (323/496) and 35% (173/496) had confirmed and probable COVID-19, respectively. The index test was positive in 255/496 (51%) patients with the primary outcome, giving a sensitivity and specificity of 51.4% (95% confidence interval [CI] 48.8 to 54.1%) and 99.5% (95% CI 99.0 to 99.8%). Sensitivity increased in those undergoing 2, 3 or 4 tests to 60.1% (95% CI 56.7 to 63.4%), 68.3% (95% CI 64.0 to 72.3%) and 77.6% (95% CI 72.7 to 81.9%), respectively. The sensitivity of the index test was 78.9% (95% CI 74.4 to 83.2%) for the secondary outcome of confirmed COVID-19 on serial testing.

**Conclusions:** In patients admitted to hospital, a single combined nasal and throat swab with RT-PCR for SARS-CoV-2 has excellent specificity, but limited diagnostic sensitivity for COVID-19. Diagnostic performance is significantly improved by repeated testing.

## Introduction

Severe acute respiratory syndrome coronavirus-2 (SARS-CoV-2) is a novel strain of coronavirus, which is responsible for the global pandemic of coronavirus disease 2019 (COVID-19). ^1,2^ Timely and accurate diagnostic testing in patients with suspected COVID-19 is essential to guide treatment and implement infection control measures to limit spread of the virus.

Reverse transcriptase-polymerase chain reaction (RT-PCR) assays on material collected by swabbing the nose and throat are the most commonly used diagnostic tests. ^3^ However, a number of reports have indicated discordance between the results of testing and clinical or radiological findings in patients with symptoms of suspected COVID-19 and increasingly clusters of asymptomatic carriers of SARS-CoV-2 are recognised. ^4-8^ As such, uncertainty remains as to the diagnostic performance of the combined nasal and throat swab to diagnose the clinical condition of COVID-19, particularly in patients presenting late following the onset of symptoms when the viral load may be lower.

Our aim was to evaluate the performance of the combined nasal and throat swab for the clinical diagnosis of COVID-19 in consecutive patients admitted to hospital with suggestive symptoms, and to determine whether there is heterogeneity across subgroups.

## Methods

### Study design

In a prospective, multi-centre, cohort study in secondary and tertiary care hospitals in Scotland, United Kingdom, we evaluated the diagnostic performance of the combined nasal and throat swab with RT-PCR for SARS-CoV-2 in consecutive patients admitted to hospital for symptoms of suspected COVID-19. The study was performed with approval of the local Research Ethics Committee and delegated Caldicott Guardian for the National Health Service (NHS) Lothian Health Board, in accordance with the Declaration of Helsinki. All data were collected from the patient record and national registries, deidentified and linked in a data repository (DataLoch ™, Edinburgh, United Kingdom) within a secure safe haven. Individual patient consent was not sought, and only summary data was extracted to minimise the risk of disclosure.

### Participants

Consecutive adult patients ≥18 years old were identified by the attending clinician using an electronic form integrated into the care pathway at the time of testing with the combined nasal and throat swab. Patients were eligible for inclusion if they were admitted to hospital with symptoms suggestive of COVID-19 and had a reportable SARS-CoV-2 RT-PCR result from material obtained through the combined swab ***(Figure 1)***. Patients were excluded if they had no symptoms and testing was performed for screening purposes only, or if they had a previous diagnosis of COVID-19.

**Figure 1.**
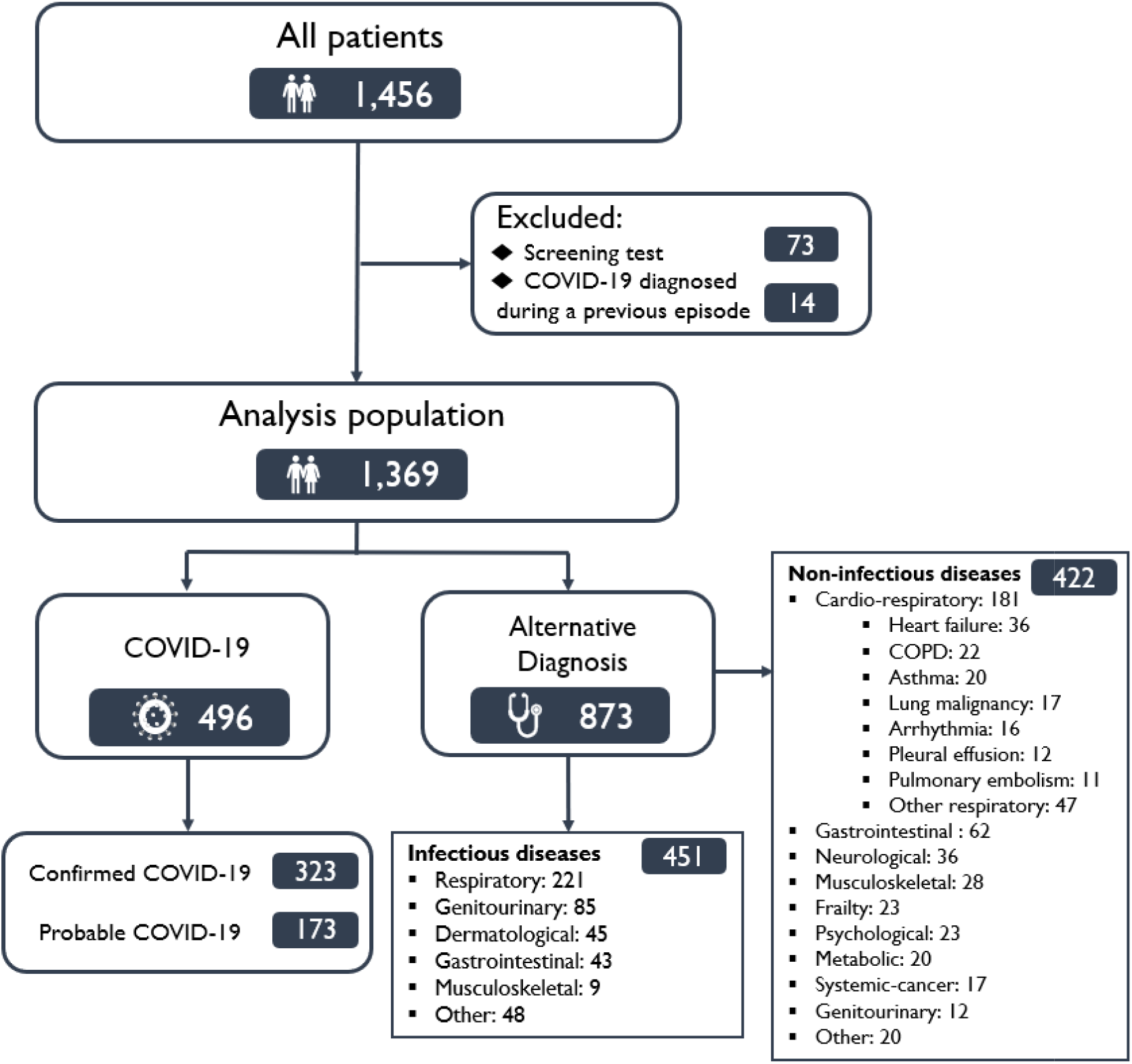
Flow diagram of study population.

### Procedures

During the study period RT-PCR for SARS-CoV-2 was performed on material obtained from a combined nasal and throat swab in patients presenting with symptoms suggestive of COVID-19 who were admitted to hospital. Repeat testing was at the discretion of the clinician responsible for care. RNA gene targets differ by manufacturer, with the tests performed here targeting one or more of the envelope (env), nucleocapsid (n), spike (s), RNA-dependent RNA polymerase (RdRp), and open reading frame-1 (ORF1) genes. The sensitivities of the tests to individual genes are comparable. ^9^ Although patients were enrolled and underwent assessment and sampling at three hospitals in the region, RT-PCR was performed in a single regional virology laboratory at the Royal Infirmary of Edinburgh.

An electronic form was embedded into the order of the combined nasal and throat swab to prospectively record information on the indication for testing, symptom type, and duration of symptoms. This form was completed by the usual care clinician at the time of testing. Data was extracted from the electronic patient record (TrakCare; InterSystems Corporation, Cambridge, MA, USA), laboratory information management system (iLaboratory, Advanced Expert Systems Medical, Derby, United Kingdom), the Scottish Morbidity Record, the Scottish Drug Dispensing Database, and the Scottish Care Information store. The Scottish Index of Multiple Deprivation (SIMD), an area-based measure of deprivation, was used to define socioeconomic status of each individual based on 31 indicators across 7 domains (income, employment, health, education, skills and training, housing, geographic access, and crime). ^10^

### Outcomes

Diagnostic performance of the index test was evaluated for a primary outcome of probable or confirmed COVID-19, and a secondary outcome of confirmed COVID-19 on serial testing. All clinical diagnoses were adjudicated by an independent, inter-disciplinary panel of clinicians using all information available within the electronic patient record, including contact history and review of all laboratory and radiological imaging performed.

The diagnosis of COVID-19 was based on the case definition proposed by the World Health Organisation. ^11^ The panel identified patients as suspected COVID-19 where they were admitted to hospital with an acute respiratory illness (fever with at least one sign or symptom of respiratory disease such as cough or shortness of breath) and had no alternative diagnosis that fully explained the clinical presentation. The panel recorded clinical, laboratory and radiological features of suspected COVID-19 without knowledge of the index and subsequent test results. Patients with parameters consistent with COVID-19 were subsequently classified as probable COVID-19 where all tests for the SARS-CoV-2 virus from any sample type were negative, or confirmed COVID-19 where any test was positive during the hospital episode, or within 7 days of the index presentation in those discharged.

For the evaluation of diagnostic performance for the primary outcome, patients were classified into the following groups: 1) *Confirmed COVID-19* in those with acute respiratory illness AND a positive test for SARS-CoV-2 (true positives); 2) *Probable COVID-19* in those with acute respiratory illness AND negative tests for SARS-CoV-2 AND no other diagnosis to explain the clinical presentation (false negatives); 3) Alternative diagnosis that fully explained their clinical presentation AND a positive test for SARS-CoV-2 (false positive); or 4) Alternative diagnosis that fully explained their clinical presentation AND negative tests for SARS-CoV-2 (true negatives).

For evaluation of diagnostic performance for the secondary outcome, patients with confirmed COVID-19 were classified as true positives, and those with probable COVID-19 were classified as true negatives rather than false negatives.

### Sample size and power

Based on data from the Hubei and Shandong provinces and Beijing, China, we anticipated that approximately 32% (126/398) of combined nasal and throat swabs performed would be positive for SARS-CoV-2. ^6^ We recognise that there may be differences in the approach to the selection of patients for testing between countries, and therefore our sample size was based on a more conservative positive test rate of 20%. We estimated that with 1,000 patients, we will have 80% power to estimate the confidence interval for a sensitivity of 90% with lower and upper intervals of 85% and 95% respectively.

### Statistical analysis

Baseline characteristics, clinical features, and laboratory results are summarised as number (percentage) or median (interquartile range) for the study population, and stratified according to the adjudicated diagnosis. Two-by-two contingency tables were constructed to compare the index test (positive or negative) in those with and without the primary and secondary outcome on serial testing (reference standard). Test sensitivity, specificity, negative predictive value (NPV) and positive predictive value (PPV) with 95% confidence intervals was determined in all participants and in pre-specified subgroups including age, sex, duration of symptoms prior to testing, fever, and respiratory tract symptoms. In patients where more than one test was performed, we report test results and compare diagnostic performance of the index test with the performance of multiple tests. All analyses were performed using R (version 3.6.1).

## Results

Between April 3 and 20, 2020, we enrolled 1,369 consecutive patients (median age 68 [interquartile range, IQR 53-80] years, 47% women) who underwent a total of 3,822 combined nasal and throat swab tests (median 2 [IQR 1-3] tests per patient) for symptoms of suspected COVID-19 (***Figure 1***). The primary outcome occurred in 36% (496/1,369), of whom 65% (323/496) and 35% (173/496) had confirmed and probable COVID-19, respectively. Of those with an alternative diagnosis (64% [873/1,269]), the most frequent diagnoses were other respiratory infections (25%, [221/873]) and non-communicable cardiorespiratory conditions (21%, [181/873]), such as heart failure, chronic obstructive pulmonary disease and asthma.

Patients with confirmed or probable COVID-19 were older than those with an alternative diagnosis (median age of 71 [57-82] *versus* 67 [53-80] years), more likely to be men (57% [281/496] *versus* 52% [450/873]) and were less likely to be from an area of deprivation (13% [65/496] *versus* 20% [174/873]). However, they had similar comorbidities and were as likely to be receiving angiotensin converting enzyme inhibitors, corticosteroids or immunosuppressants at presentation ***(Table 1 and eTable 1 in the Supplement)***.

**Table 1.**
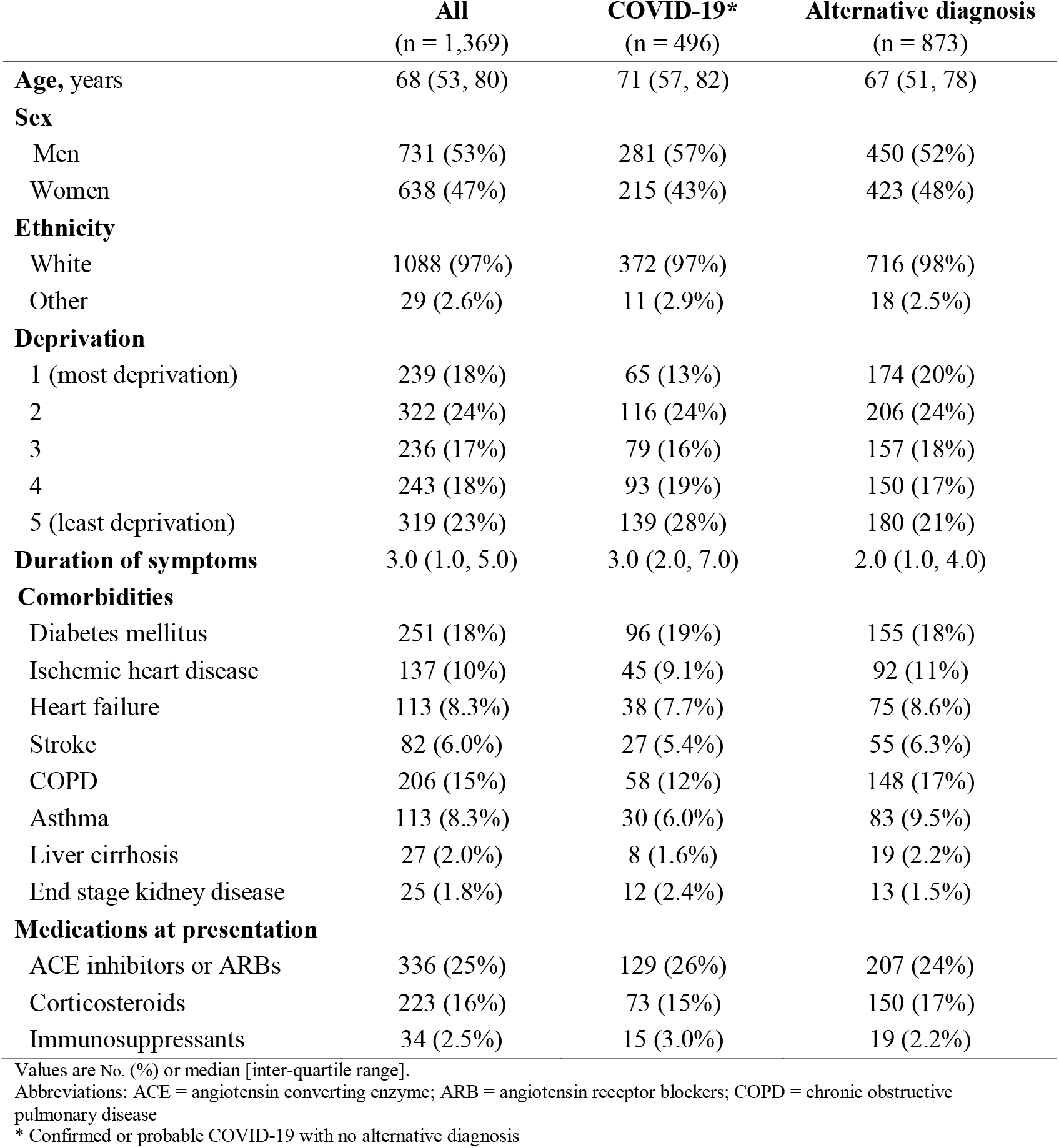
Baseline characteristics of patients undergoing testing with suspected COVID-19

Compared to patients with an alternative diagnosis, those with confirmed or probable COVID-19 had a lower lymphocyte and neutrophil count (median 1.14 x10 ^9^/L *versus* 1.44 x10 ^9^/L and 5.8 x10 ^9^/L *versus* 6.4 x10 ^9^/L, respectively), but a higher C-reactive protein concentration (52 mg/dL *versus* 22 mg/dL) at presentation ***(eTable 2 and 3 in the Supplement)***. In patients with probable or confirmed COVID-19, compared to those with an alternative diagnosis, some symptoms and signs were more common, including fever on presentation (65% [322/496] *versus* 42% [369/873]), upper and lower respiratory tract symptoms (13% [63/496] *versus* 10% [78/873]; 85% [419/496] *versus* 54% [473/873], respectively), and systemic symptoms (52% [257/496] *versus* 28% [244/873]). In contrast, there were no differences in the frequency of neurological (24% [118/496] *versus* 22% [191/873]) or gastrointestinal symptoms (22% [107/496] *versus* 24% [209/873]) ***(Figure 2a and eTable 4 in the Supplement)***. Patients with probable or confirmed COVID-19 were six-times more likely to have radiological signs of infection on chest imaging than those with an alternative diagnosis (64% [316/496] versus 11% [93/873]). Patients with confirmed COVID-19 had similar symptoms as those with probable COVID-19 ***(Figure 2b and eTable 5 in the Supplement)***, but were more likely to have lymphopenia, systemic inflammation and radiological signs of infection.

**Figure 2.**
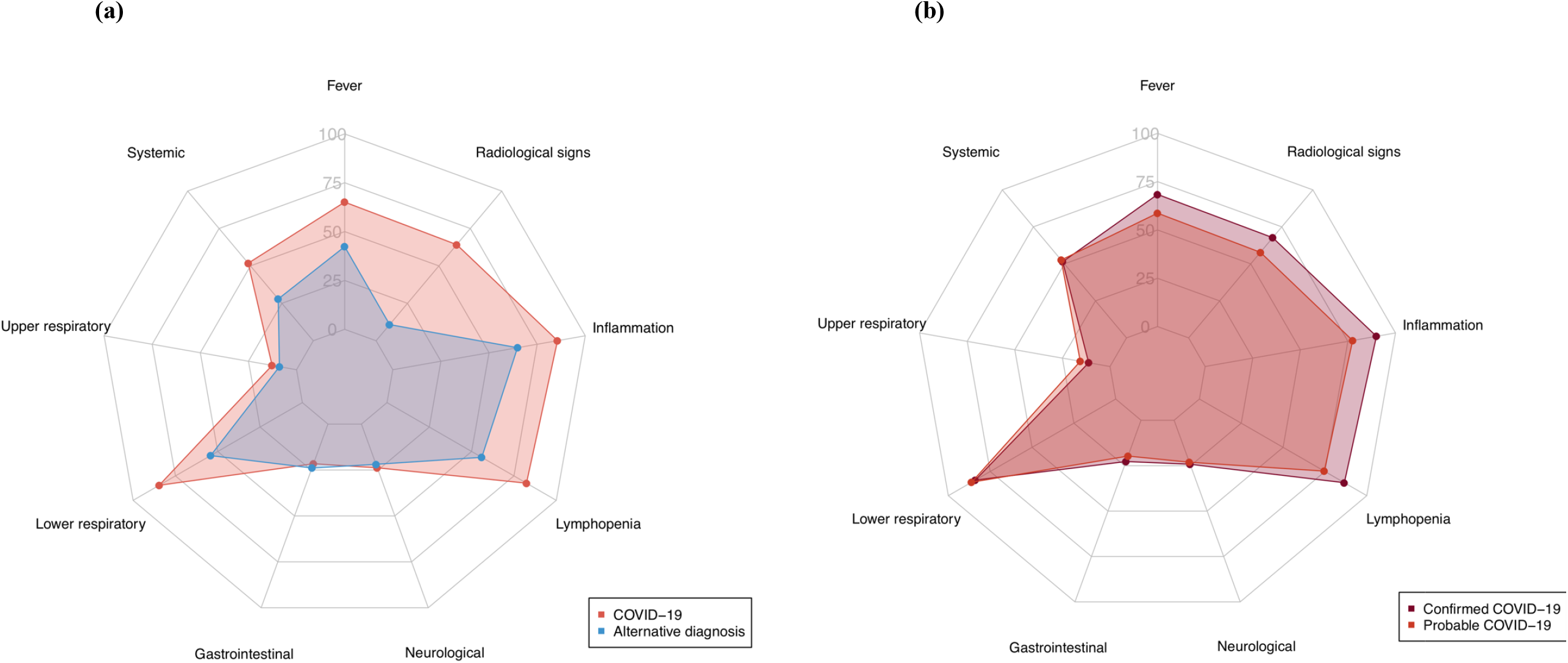
Radar plot of the clinical features of patients with (a) an adjudicated diagnosis of confirmed or probable COVID-19 and those with an alternative diagnosis, and (b) confirmed COVID-19 and those with probable COVID-19 *The following features used to adjudicate the diagnosis are illustrated: fever, systemic symptoms (e*.*g. myalgia, fatigue, and arthralgia), upper respiratory tract symptoms, lower respiratory tract symptoms, gastrointestinal symptoms, neurological symptoms, lymphopenia, systemic inflammation, radiological features of infection*.

The index test was positive in 255/496 (51%) patients with the primary outcome ***(eTable 2 in the Supplement)***, giving a sensitivity, specificity, negative predictive value and positive predictive value of 51.4% (95% confidence interval [CI] 48.8 to 54.1%), 99.5% (95% CI 99.0 to 99.8%), 78.3% (95% CI 76.0 to 80.4%) and 98.5% (95% CI 97.7 to 99.0%), respectively ***(eTable 6 in the Supplement)***. Sensitivity was lower in patients from areas with the greatest deprivation (32.6% [95% CI 21.6 to 43.9%] *versus* 53.9% [95% CI 45.7 to 62.1%], quintile with the most deprivation compared to the quintile with the least deprivation). Otherwise, sensitivity remained consistent across other patient demographic factors, comorbidities and symptoms ***(Figure 3a)***. The negative predictive value of the index test was consistent across patient demographics and comorbidities, but was lower in those with fever (70.3% [95% CI 66.3 to 74.2%]), lower respiratory symptoms (69.5% [95% CI 66.1 to 73.0%]) and those with a longer duration of symptoms prior to hospital admission (68.7% [95% CI 63.2 to 74.1%] *versus* 87.1% [95% CI 83.3 to 90.7%] for patients presenting ≥4 days after symptom onset compared to those presenting within one day) ***(Figure 3b)***.

**Figure 3.**
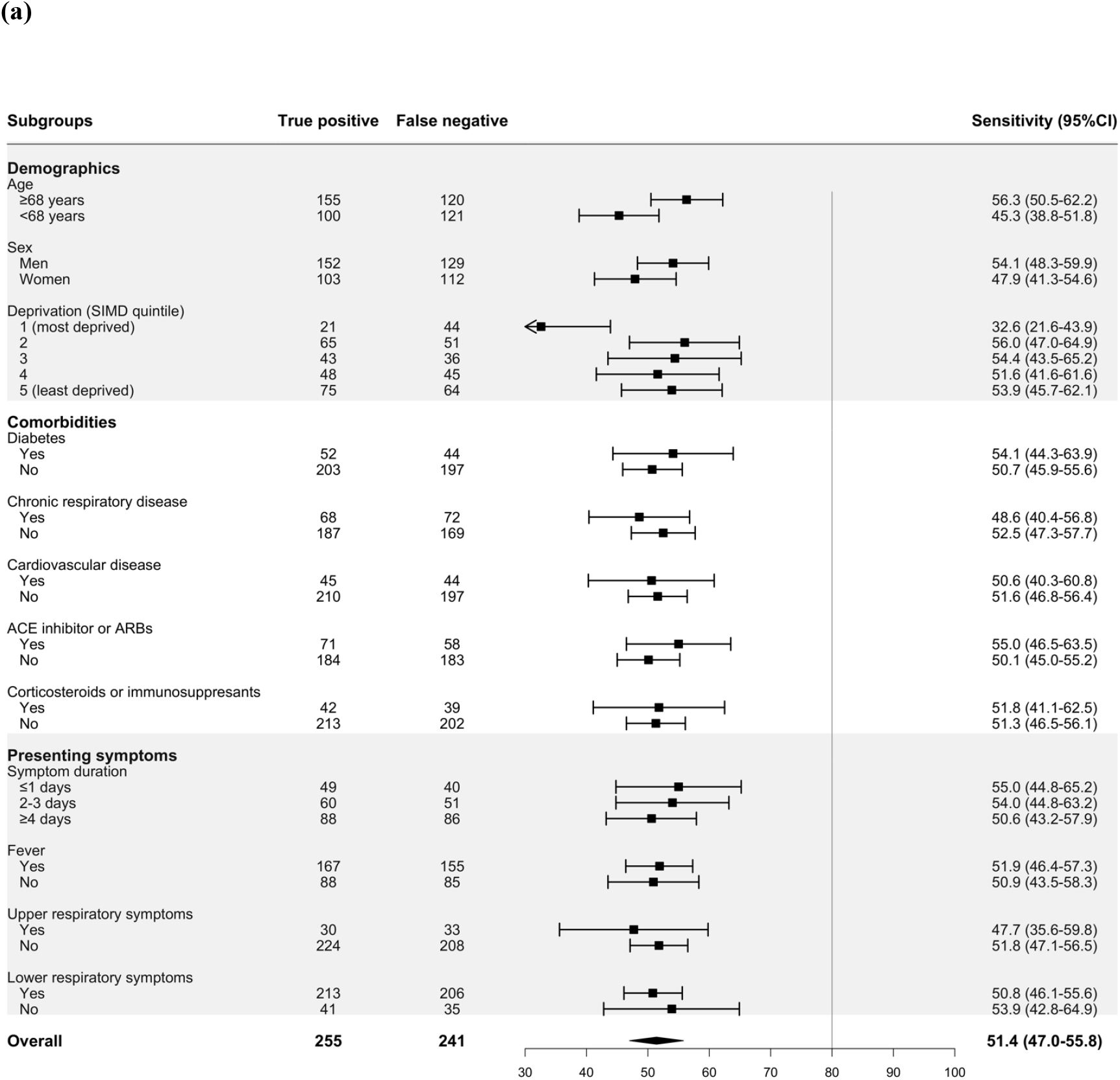
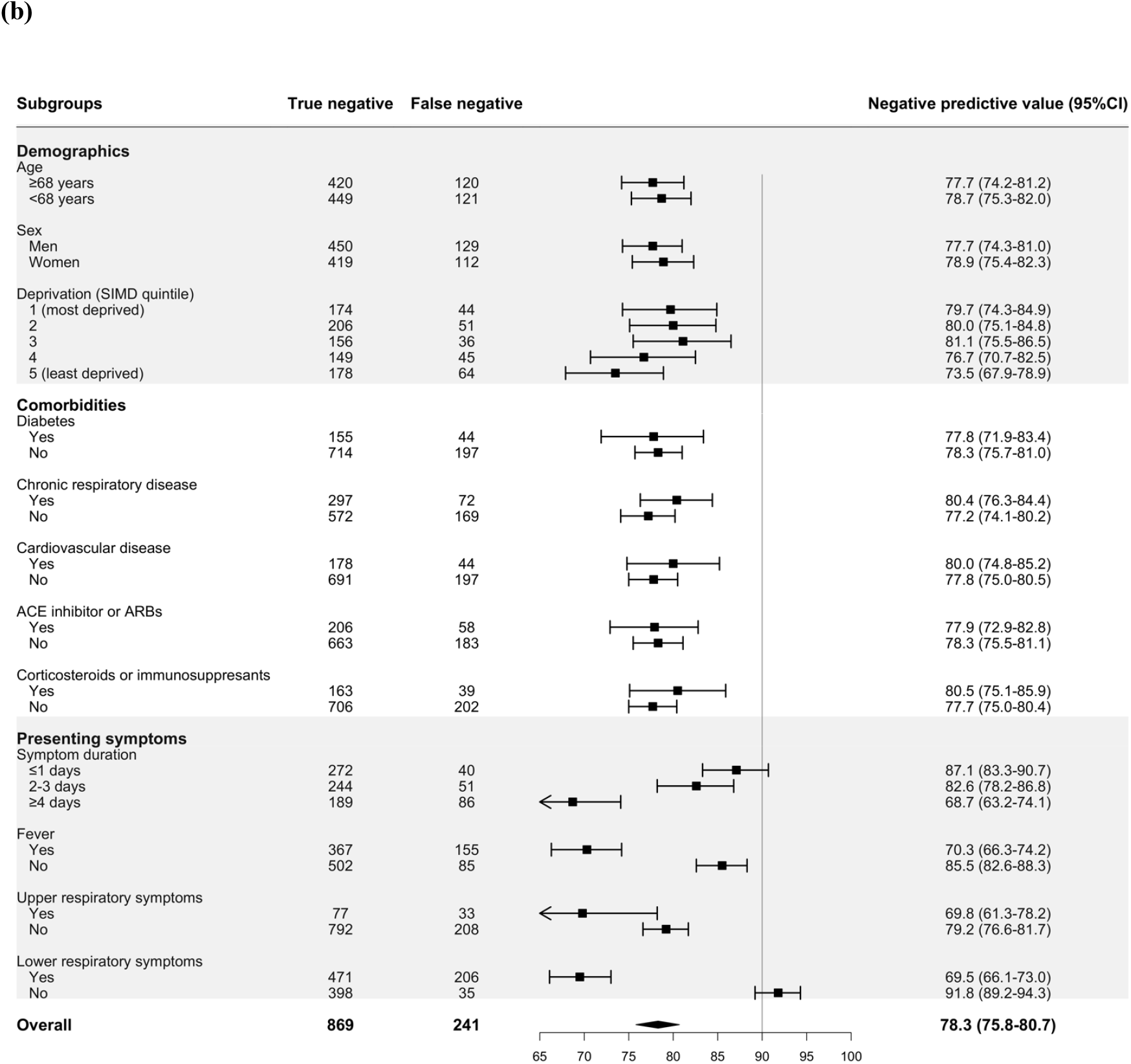
Forest plot of the (a) sensitivity and (b) negative predictive value of the index combined nasal and throat swab for a diagnosis of confirmed or probable COVID-19 stratified by subgroups.

The majority of patients underwent serial testing (59.5%, 815/1,369) with 22.6% (310/1,369) undergoing 4 or more serial tests ***(eFigure 1 and 2, and eTable 6 in the Supplement)***. Of those with confirmed COVID-19, patients with a negative index test result underwent more serial testing than those with a positive index test result (95.6% [65/68] *versus* 65.9% [168/255] with median test per patient of 4 [2-6] *versus* 2 [1-5] respectively). The median time between first and second tests was shorter in patients with confirmed COVID-19 who had a negative index test result and those with probable COVID-19 (1.7 [0.8-10.9] days and 1.2 [0.9-4.6] days respectively) compared to patients with confirmed COVID-19 who had a positive index test result and those with an alternative diagnosis (6.8 [4.0-8.6] days and 6.1 [1.1-21.8] days respectively). Sensitivity for the primary outcome increased in those undergoing 2, 3 or 4 serial tests to 60.1% (95% CI 56.7 to 63.4%), 68.3% (95% CI 64.0 to 72.3%) and 77.6% (95% CI 72.7 to 81.9%), respectively ***(Figure 4 and eTable 7 in the Supplement)***. The negative predictive value increased more modestly on serial testing; from 78.3% (76.0 to 80.4%) for the index test to 79.8% (75.0 to 83.9%) in those undergoing 4 serial tests. These observations persisted in a sensitivity analysis restricted to patients who underwent at least 4 tests and for the secondary outcome ***(Figure 4, eFigure 3 and 4, and eTable 8 in the Supplement)***.

**Figure 4.**
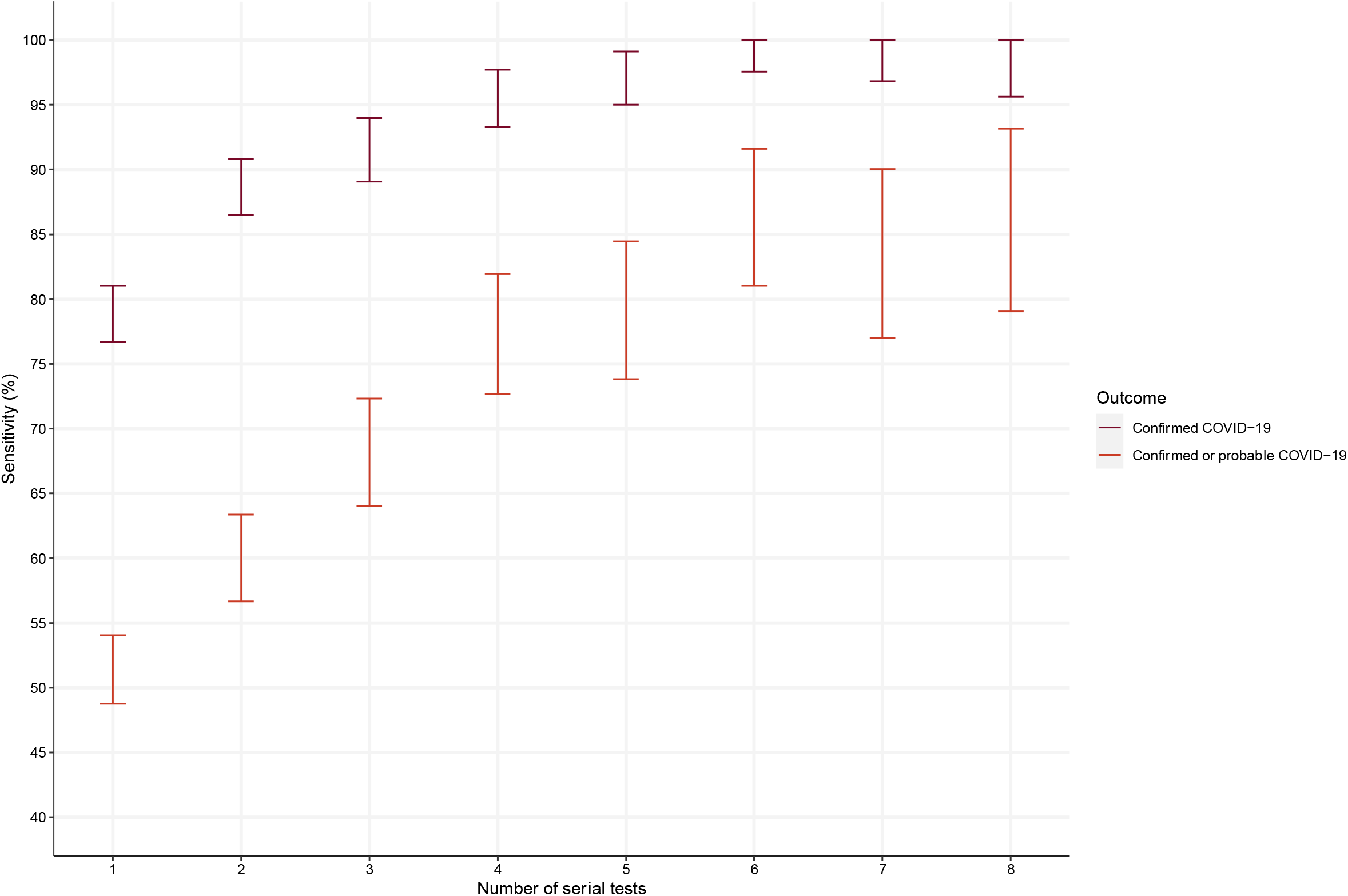
Sensitivity of serial testing using the combined nasal and throat swab for the primary (confirmed or probable COVID-19) and secondary (confirmed COVID-19) outcome.

Sensitivity of the index test for the secondary outcome of a diagnosis of confirmed COVID-19 on serial testing was 78.9% (95% CI 74.4 to 83.2%) ***(eFigure 5a in the Supplement)***. There was no significant heterogeneity in sensitivity across patient demographics, comorbidities or presenting symptoms. The negative predictive value of the index test was 93.8% (95% CI 92.4 to 95.2%) for a diagnosis of confirmed COVID-19 on serial testing ***(eFigure 5b in the Supplement)***. The negative predictive value remained consistent across patient demographics and comorbidities, but was lower in patients who presented with a fever (89.6% [95% CI 86.9 to 92.1%]) and those who had lower respiratory symptoms (91.5% [95% CI 89.4 to 93.6%]).

## Discussion

In this prospective, multi-centre, cohort study, we evaluated the diagnostic performance of the combined nasal and throat swab with RT-PCR for SARS-CoV-2 in consecutive patients admitted to hospital with symptoms of suspected COVID-19. We report a number of potentially important findings. First, a single test had excellent specificity, but limited sensitivity for an adjudicated diagnosis of probable or confirmed COVID-19. Second, the sensitivity of the index test was higher for patients with confirmed COVID-19 on serial testing, but still missed 1 in 5 patients with the diagnosis. Third, diagnostic performance was similar in most patient subgroups, but the sensitivity was lower in those from more deprived areas, and the negative predictive value was lower in those presenting later following the onset of symptoms, and in those with fever or lower respiratory symptoms. Finally, we observed a significant improvement in diagnostic sensitivity with repeated testing on up to four occasions.

Our study has several strengths. This was a prospective, multi-centre study that was adequately powered to evaluate the diagnostic performance of the combined nasal and throat swab. Patients were enrolled using an electronic form embedded within clinical care across all secondary and tertiary hospitals in the region. This permitted us to include all consecutive patients who underwent testing for symptoms that were considered to be suggestive of COVID-19 by their usual care clinician minimizing selection bias and ensuring our findings are representative of all hospitalised patients across the region. The diagnosis was adjudicated by a multidisciplinary panel of clinicians from a range of specialities involved in the care of patients with COVID-19, including infectious disease, emergency medicine, general medicine and geriatric medicine, and therefore our findings are relevant to clinical practice across secondary and tertiary care settings. To our knowledge this is the first evaluation of the diagnostic performance of the combined nasal and throat swab for a clinical diagnosis of COVID-19. The evaluation of the combined nasal and throat swab is particularly important since this is the most widely used diagnostic modality to identify or exclude SARS-CoV-2 infection. Although the RT-PCR test for SARS-CoV-2 has excellent *in vitro* analytical performance under carefully controlled laboratory conditions, ^9^ diagnostic performance in clinical practice can vary due to multiple other factors, such as the site and quality of sampling, stage of disease, and viral multiplication or clearance. ^3,6,12^ Previous studies have reported diagnostic sensitivities ranging from 50% to 100% for the combined nasal and throat swab with a meta-analysis reporting a pooled meta-estimate of 89% (95% CI 81 to 94%). ^4,6,13-16^ However, these studies have been performed in relatively small, selected patient cohorts, which limit the generalisability of study findings across the breadth of patients presenting to hospitals with suspected COVID-19. Furthermore, the reference standard used in these studies was either radiological findings on chest computed tomography, or the results of subsequent RT-PCR tests, which inevitably leads to an increase in the estimated diagnostic sensitivity. Indeed, when our reference standard was restricted to confirmed COVID-19 on serial testing, diagnostic sensitivity increased from 51.4% to 78.9%.

In subgroup analyses, the diagnostic performance was similar in older patients, those with diabetes, known respiratory or cardiovascular disease. These findings are reassuring since these patient subgroups have been identified as those at the highest risk of death from COVID-19. ^17-19^ However, we observed that diagnostic sensitivity was lower in patients from the most deprived areas. This may reflect differences in the clinicians approach to testing by deprivation, rather than a consequence of deprivation itself, but this requires further evaluation. Furthermore, we observed that negative predictive value was lower in those who presented late in the course of their illness, and those with symptoms of fever and lower respiratory tract symptoms. This is consistent with virological assessments of patients hospitalised with COVID-19 which showed that viral RNA shedding from the upper respiratory tract is typically highest at the onset of symptoms, but subsequently declines as the disease progresses. ^12,20^ The lower negative predictive value in those with typical clinical symptoms is likely to reflect a higher pre-test probability for COVID-19 in these patients.

Our findings have potentially important implications for the use and interpretation of this test in clinical practice. Whilst the diagnosis of COVID-19 is still largely reliant on RT-PCR on material collected on nose and throat swabs, clinicians need to be aware of the strengths and limitations of testing when making decisions on the placement of patients within hospital settings and discharge planning. In our study, the most conservative estimate of diagnostic sensitivity was 51%, where the index test was negative in 1 in 2 patients with the primary outcome of probable or confirmed COVID-19. Our most optimistic estimate of diagnostic sensitivity was 79%, where the index test was negative in 1 in 5 patients with the secondary outcome of confirmed COVID-19. Whilst, the former may underestimate performance, the latter is certainly an overestimate due to circular reasoning, whereby the test under evaluation is an essential component of the reference standard.

In this consecutive series of hospitalised patients where testing was performed for symptoms at the discretion of the usual care clinician, our multi-disciplinary panel diagnosed probable COVID-19 in 12.6% of patients. The panel included representation from a broad range of medical specialities involved in the assessment of these patients, and therefore their judgment is likely to be representative of clinical practice. Our approach aims to provide insights into the performance of the test as it is applied in clinical practice. Interestingly patients with probable COVID-19 or confirmed COVID-19 and a negative index test were more likely to be retested within the next 24-48 hours. This likely reflects the usual care clinician’s uncertainty when interpreting a negative test. Indeed for both the primary and secondary outcome, diagnostic performance improved significantly with up to four serial tests, and this observation could inform our approach to serial testing in practice with implications for patient flow and management of hospitalised patients. Our findings also have implications when defining the reference standard for studies evaluating the performance of point of care ^21^ and laboratory antibody tests ^22^ to determine those with and without prior infection. Future research should evaluate performance in patients undergoing testing in the community, and determine whether performance can be improved by incorporating a measure of sampling efficacy using epithelial cell counts ^23^, or whether the diagnostic yield is higher in other sample types, such as saliva or sputum.

We acknowledge our study has several limitations. We did not mandate the number of serial tests as all diagnostic testing was performed at the discretion of the treating clinician. Therefore those with a negative index test undergoing serial testing are likely to have had a higher pre-test probability than those undergoing a single test. Nevertheless, it is reassuring that a similar increase in diagnostic performance with serial testing was observed in a sensitivity analysis restricted to those patients who had a complete series of at least four tests. Further studies with systematic sampling in consecutive patients are required to validate this observation. In the absence of an independent gold standard test for the diagnosis of COVID-19, our diagnostic evaluation was based on clinical review of all tests ordered by the usual care clinician. As a comprehensive panel of respiratory pathogens was not requested in all patients, it is likely we have misclassified some patients as COVID-19 who had other viral or bacterial infections. We may have overestimated the diagnosis of COVID-19 given the study was performed during the peak of a pandemic. However, we reviewed all available clinical investigations including all laboratory and imaging findings, and only defined patients with suspected COVID-19 where there was no alternative diagnosis. Furthermore, the clinical features of those with probable or confirmed COVID-19 were identical, suggesting no systematic bias was introduced during the adjudication.

In conclusion, a single combined nasal and throat swab with RT-PCR for SARS-CoV-2 has excellent specificity, but limited diagnostic sensitivity for the clinical diagnosis of COVID-19. Diagnostic performance is significantly improved by repeated testing.

## DataLoch COVID-19 Collaborators

### Clinical Lead

Atul Anand

### Programme Lead

Kathy Harrison

### DataLoch Technical and Management Group

Catherine Stables, Ally Hume, David Homan, Catriona Waugh, Jilly McKay, Chris Duncan, Ronnie Harkess

### DataLoch Leadership Group

Kathy Harrison, Michael Gray, Colan Mahaffey, Pamela Linksted, Atul Anand, Anoop SV Shah, Rob Baxter, Peter Cairns, Nicola Rigglesford, Martin Egan, Nicholas L Mills

### Infectious Diseases Unit

Daniella A Ross, Claire L Mackintosh, Oliver Koch, Kate Templeton, Meghan R Perry

### COVID-19 Clinical Data Review

Daniella A Ross, Anda Bularga, Hannah MM Preston, Thomas J McCormick, Arjuna A Sivakumaran, Kathryn AW Knight, Rosie Callender, Anna K Jamieson, Jonathan Wubetu, John P Kelly, Zaina Sharif, Ha Bao Trung Le, Jason Yang, Arun Parajuli, Ed Whittaker, Oscar CN Maltby, Sarah H Goodwin, Louisa R Cary, Emma K Watson, Thomas H Clouston, Julia Guerrero Enriquez, XinYi Ng

### COVID-19 Adjudication Panel

Kuan Ken Lee, Daniella A Ross, Anda Bularga, Andrew R Chapman, Yvonne K McFarlane, Kate H Regan, Richard P Biggers, John P Kelly, Kathryn AW Knight, Hannah MM Preston, Thomas J McCormick, Anoop SV Shah, Atul Anand, Meghan R Perry, Nicholas L Mills

### NHS Lothian eHealth and Lothian Analytical Services

Alistair Stewart, Alastair Thomson, Chris Duncan, Daniella Ene, Hazel Neilson, Juergen Caris, Maria McMenemy, Nazir Lone, Nicola Rigglesford, Paul Schofield, Sophie McCall, Stephen Young, Tracey McKinley, Tracey Rapson

## Data Availability

All data were collected from the patient record and national registries, deidentified and linked in a data repository (DataLochTM, Edinburgh, United Kingdom) within a secure safe haven.

## Acknowledgements

This study was funded by a British Heart Foundation (BHF) Research Excellence Award (RE/18/6134217). DataLoch ™ is funded by the University of Edinburgh, and the UK and Scottish Governments as part of the Data Driven Innovation in Health & Social Care programme. AA is supported by a Clinical Lectureship from the Chief Scientist Office (PCL/18/05). KL and NLM are supported by a Clinical Research Training Fellowship (FS/18/25/33454) and the Butler Senior Clinical Research Fellowship (FS/16/14/32023) from the British Heart Foundation, respectively. DD is supported by the Medical Research Council (MR/N013166/1). ARC is supported by a Starter Grant for Clinical Lecturers from the Academy of Medical Sciences (SGL021\1075).

## Author Contributions

KKL, AA, ASVS and NLM conceived the study and its design. KKL, DR, AB, ARC, ASVS, AA, MP, and NLM acquired the data. KKL and DD performed the analysis. KKL, DD, DR, AB, CM, OK, IJ, SJ, ARC, ASVS, AA, MP, and NLM interpreted the data. KKL and NLM drafted the manuscript. All authors revised the manuscript critically for important intellectual content and provided their final approval of the version to be published. All authors are accountable for the work.

