## Supplementary material for "Diagnostic performance of the combined nasal and throat swab in patients admitted to hospital with suspected COVID-19": Data Supplement

**Diagnostic performance of the combined**

**nasal and throat swab in patients**

**admitted to hospital with suspected COVID-19**

Kuan Ken Lee, M.D.,^1^ Dimitrios Doudesis, M.Sc.,^1, 2^ Daniella A. Ross, M.D.,^3^

Anda Bularga, M.D.,^1^ Claire L. MacKintosh, M.D. , Ph.D.,^3^ Oliver Koch, M.D., Ph.D.,^3^

Ingolfur Johannessen, M.D., Ph.D.,^4^ Kate Templeton, Ph.D., FRCPath.,^4^

Sara Jenks, M.D., FRCPath.,^5^ Andrew R. Chapman, M.D., Ph.D.,^1^

Anoop S.V. Shah, M.D., Ph.D.,^1,6,7^ Atul Anand, M.D., Ph.D.,^1^

Meghan R. Perry, M.D., Ph.D.,^3^ Nicholas L. Mills, M.D., Ph.D.^1, 2^

*on behalf of the DataLoch COVID-19 Collaboration**

^1^ BHF Centre for Cardiovascular Science, University of Edinburgh, Edinburgh, UK.

^2^ Usher Institute, University of Edinburgh, Edinburgh, UK.

^3^ Regional Infectious Disease Unit, Western General Hospital, Edinburgh, UK.

^4^ Department of Clinical Virology, Royal Infirmary of Edinburgh, Edinburgh, UK

^5^ Department of Clinical Biochemistry, Royal Infirmary of Edinburgh, Edinburgh, UK.

^6^ Department of Non-communicable Disease Epidemiology, London School of Hygiene and Tropical Medicine, London, UK

^7^ Department of Cardiology, Imperial College Healthcare NHS Trust, London, UK

**Corresponding Author:**

Professor Nicholas L Mills

BHF/University Centre for Cardiovascular Science

The University of Edinburgh

Edinburgh EH16 4SA

United Kingdom

**eTables:**	8

**eFigures:**	5

**eTable 1.** Baseline characteristics of patients stratified according to whether the diagnosis of COVID-19 was confirmed or probable

|  | **All**  **COVID-19**  (n = 496) | **Confirmed COVID-19**  (n = 323) | **Probable**  **COVID-19**  (n = 173) |
| --- | --- | --- | --- |
| **Age,** years | 71 (57, 82) | 73 (59, 83) | 66 (52, 78) |
| **Sex** |  |  |  |
| Men | 281 (57%) | 193 (60%) | 88 (51%) |
| Women | 215 (43%) | 130 (40%) | 85 (49%) |
| **Ethnicity** |  |  |  |
| White | 372 (97%) | 238 (96%) | 134 (99%) |
| Other | 11 (2.9%) | 10 (4.0%) | 1 (0.7%) |
| **Deprivation** |  |  |  |
| 1 (most deprived) | 65 (13%) | 32 (10%) | 33 (19%) |
| 2 | 116 (24%) | 81 (25%) | 35 (20%) |
| 3 | 79 (16%) | 51 (16%) | 28 (16%) |
| 4 | 93 (19%) | 66 (21%) | 27 (16%) |
| 5 (least deprived) | 139 (28%) | 89 (28%) | 50 (29%) |
| **Duration of symptoms** | 3.0 (2.0, 7.0) | 3.0 (2.0, 7.0) | 4.0 (2.0, 10.0) |
| **Comorbidities** |  |  |  |
| Diabetes mellitus | 96 (19%) | 65 (20%) | 31 (18%) |
| Ischemic heart disease | 45 (9.1%) | 24 (7.4%) | 21 (12%) |
| Heart failure | 38 (7.7%) | 27 (8.4%) | 11 (6.4%) |
| Stroke | 27 (5.4%) | 15 (4.6%) | 12 (6.9%) |
| COPD | 58 (12%) | 33 (10%) | 25 (14%) |
| Asthma | 30 (6.0%) | 22 (6.8%) | 8 (4.6%) |
| Liver cirrhosis | 8 (1.6%) | <5 | <5 |
| **Medications at presentation** |  |  |  |
| ACE inhibitors or ARBs | 129 (26%) | 89 (28%) | 40 (23%) |
| Corticosteroids | 73 (15%) | 48 (15%) | 25 (14%) |
| Immunosuppressants | 15 (3.0%) | 9 (2.8%) | 6 (3.5%) |

Values are No. (%) or median [inter-quartile range].

Abbreviations: ACE = angiotensin converting enzyme; ARB = angiotensin receptor blockers; COPD = chronic obstructive pulmonary disease

**eTable 2.** Virology, laboratory tests at presentation with suspected COVID-19

|  | **All**  (n = 1,369) | **COVID-19***  (n = 496) | **Alternative diagnosis**  (n = 873) | **P-value** |
| --- | --- | --- | --- | --- |
| Positive index PCR test | 259 (19%) | 255 (51%) | 4 (0.5%)^†^ | <0.001 |
| White cell count, x10^9^/L | 8.9 (6.5, 12.5) | 8.1 (6.0, 11.8) | 9.4 (6.9, 13.0) | <0.001 |
| Lymphocyte count, x10^9^/L | 1.32 (0.87, 1.89) | 1.14 (0.78, 1.61) | 1.44 (0.94, 2.03) | <0.001 |
| Neutrophil count, x10^9^/L | 6.2 (4.2, 9.4) | 5.8 (4.0, 8.9) | 6.4 (4.4, 9.9) | <0.001 |
| C-reactive protein, mg/dL | 32 (7, 105) | 52 (13, 123) | 22 (6, 94) | <0.001 |
| ALT, IU/L | 20 (13, 33) | 22 (14, 36) | 18 (13, 32) | <0.001 |
| Impaired renal function | 399 (30%) | 158 (32%) | 241 (28%) | 0.11 |

Values are No. (%) or median [inter-quartile range].

Abbreviations: ALT = alanine transaminase; PCR = polymerase chain reaction

Impaired renal function = estimated glomerular filtration rate of <60 mL/min

* Confirmed or probable COVID-19 with no alternative diagnosis

† Four patients were adjudicated to be asymptomatic carriers of SARS-CoV-2

**eTable 3.** Virology, laboratory tests at presentation stratified according to whether the diagnosis of COVID-19 was confirmed or probable

|  | **All**  **COVID-19**  (n = 496) | **Confirmed COVID-19**  (n = 323) | **Probable**  **COVID-19**  (n = 173) | **P-value** |
| --- | --- | --- | --- | --- |
| Positive index PCR test | 255 (51%) | 255 (79%) | 0 (0%) | <0.001 |
| White cell count, x10^9^/L | 8.1 (6.0, 11.8) | 7.8 (5.8, 11.2) | 9.1 (6.4, 12.3) | 0.016 |
| Lymphocyte count, x10^9^/L | 1.14 (0.78, 1.61) | 1.08 (0.72, 1.52) | 1.22 (0.90, 1.79) | 0.003 |
| Neutrophil count, x10^9^/L | 5.8 (4.0, 8.9) | 5.8 (3.9, 8.5) | 5.9 (4.2, 9.5) | 0.150 |
| C-reactive protein, mg/dL | 52 (13, 123) | 58 (18, 128) | 40 (8, 105) | 0.005 |
| ALT, IU/L | 22 (14, 36) | 22 (14, 36) | 21 (15, 34) | 0.530 |
| Impaired renal function | 158 (32%) | 110 (34%) | 48 (28%) | 0.180 |

Values are No. (%) or median [inter-quartile range].

Abbreviations: ALT = alanine transaminase; PCR = polymerase chain reaction

Impaired renal function = estimated glomerular filtration rate of <60 mL/min

**eTable 4.** Clinical features of patients undergoing testing with suspected COVID-19

| **Variable** | **All**  (n = 1369) | **COVID-19***  (n = 496) | **Alternative diagnosis**  (n = 873) | **P-value** |
| --- | --- | --- | --- | --- |
| Fever | 691 (51%) | 322 (65%) | 369 (42%) | <0.001 |
| Upper respiratory tract symptoms | 141 (10%) | 63 (13%) | 78 (8.9%) | 0.034 |
| Lower respiratory tract symptoms | 892 (65%) | 419 (85%) | 473 (54%) | <0.001 |
| Systemic symptoms | 501 (37%) | 257 (52%) | 244 (28%) | <0.001 |
| Neurological symptoms | 309 (23%) | 118 (24%) | 191 (22%) | 0.450 |
| Gastrointestinal symptoms | 316 (23%) | 107 (22%) | 209 (24%) | 0.360 |
| Lymphopenia | 894 (65%) | 406 (82%) | 488 (56%) | <0.001 |
| Inflammation | 986 (72%) | 421 (86%) | 565 (65%) | <0.001 |
| Radiological signs | 409 (30%) | 316 (64%) | 93 (11%) | <0.001 |

Values are No. (%).

* Confirmed or probable COVID-19 with no alternative diagnosis

### eTable 5. Phenotype of patients undergoing testing with suspected COVID-19 stratified according to whether the diagnosis of COVID-19 was confirmed or probable

| **Variable** | **All**  **COVID-19**  (n = 496) | **Confirmed COVID-19**  (n = 323) | **Probable**  **COVID-19**  (n = 173) | **p-value** |
| --- | --- | --- | --- | --- |
| Fever | 322 (65%) | 221 (68%) | 101 (59%) | 0.040 |
| Upper respiratory tract symptoms | 63 (13%) | 36 (11%) | 27 (16%) | 0.200 |
| Lower respiratory tract symptoms | 419 (85%) | 270 (84%) | 149 (86%) | 0.590 |
| Systemic symptoms | 257 (52%) | 166 (51%) | 91 (53%) | 0.870 |
| Neurological symptoms | 118 (24%) | 78 (24%) | 40 (23%) | 0.870 |
| Gastrointestinal symptoms | 107 (22%) | 73 (23%) | 34 (20%) | 0.510 |
| Lymphopenia | 406 (82%) | 277 (87%) | 129 (75%) | 0.001 |
| Inflammation | 421 (86%) | 287 (90%) | 134 (77%) | <0.001 |
| Radiological signs | 316 (64%) | 216 (68%) | 100 (58%) | 0.037 |

Values are No. (%).

**eTable 6.** Use of serial testing in patients with suspected COVID-19

|  | **All**  **n=1369** | **Confirmed or probable COVID-19**  **n=496** | **Confirmed COVID-19** | | **Probable COVID-19**  **n=173** | **Alternative diagnosis**  **n=873** |
| --- | --- | --- | --- | --- | --- | --- |
|  |  |  | Index test positive  n=255 | Index test negative  n=68 |  |  |
| Serial testing | 815 (59.5) | 353 (71.2) | 168 (65.9) | 65 (95.6) | 120 (69.4) | 462 (52.9) |
| Number of tests per patient | 2 [1-3] | 2 [1-4] | 2 [1-5] | 4 [2-6] | 2 [1-3] | 2 [1-3] |
| Days between first and second serial test | 5.2 [1.1-15.0] | 4.6 [1.0-8.1] | 6.8 [4.0-8.6] | 1.7 [0.8-10.9] | 1.2 [0.9-4.6] | 6.1 [1.1-21.8] |

Values are No. (%) or median [IQR].

**eTable 7.** Diagnostic performance of the index and serial combined nasal and throat swab for the primary outcome of a diagnosis of confirmed or probable COVID-19

| **Serial tests** | **True negative** | **False negative** | **True positive** | **False positive** | **Sensitivity**  **(95% CI)** | **Negative predictive value**  **(95% CI)** | **Positive predictive value**  **(95% CI)** | **Specificity**  **(95% CI)** |
| --- | --- | --- | --- | --- | --- | --- | --- | --- |
| 1 (index test) | 868 | 241 | 255 | 4 | 51.4 (48.8-54.1) | 78.3 (76.0-80.4) | 98.5 (97.7-99) | 99.5 (99.0-99.8) |
| 2 | 459 | 141 | 212 | 5 | 60.1 (56.7-63.4) | 76.5 (73.5-79.3) | 97.7 (96.4-98.5) | 98.9 (98.0-99.4) |
| 3 | 235 | 76 | 164 | 7 | 68.3 (64.0-72.3) | 75.6 (71.5-79.2) | 95.9 (93.7-97.3) | 97.1 (95.2-98.3) |
| 4 | 142 | 36 | 125 | 7 | 77.6 (72.7-81.9) | 79.8 (75.0-83.9) | 94.7 (91.6-96.7) | 95.3 (92.3-97.2) |
| 5 | 94 | 24 | 94 | 6 | 79.7 (73.8-84.5) | 79.7 (73.8-84.5) | 94.0 (90.0-96.5) | 94.0 (90.0-96.5) |
| 6 | 62 | 11 | 75 | 6 | 87.2 (81.0-91.6) | 84.9 (78.4-89.7) | 92.6 (87.3-95.8) | 91.2 (85.6-94.7) |
| 7 | 46 | 10 | 55 | 6 | 84.6 (77.0-90.0) | 82.1 (74.2-88.0) | 90.2 (83.4-94.3) | 88.5 (81.4-93.1) |
| 8 | 31 | 6 | 43 | 4 | 87.8 (79.1-93.2) | 83.8 (74.5-90.2) | 91.5 (83.6-95.8) | 88.6 (80.0-93.7) |

**eTable 8.** Diagnostic performance of the index and serial combined nasal and throat swab for the secondary outcome of a diagnosis of confirmed COVID-19 on serial testing

| **Serial tests** | **True negative** | **False negative** | **True positive** | **False positive** | **Sensitivity**  **(95% CI)** | **Negative predictive value**  **(95% CI)** | **Positive predictive value**  **(95% CI)** | **Specificity**  **(95% CI)** |
| --- | --- | --- | --- | --- | --- | --- | --- | --- |
| 1 (index test) | 1041 | 68 | 255 | 4 | 78.9 (76.7-81.0) | 93.9 (92.5-95.0) | 98.5 (97.7-99.0) | 99.6 (99.1-99.8) |
| 2 | 574 | 26 | 207 | 10 | 88.8 (86.5-90.8) | 95.7 (94.0-96.9) | 95.4 (93.7-96.6) | 98.3 (97.1-99.0) |
| 3 | 297 | 14 | 158 | 13 | 91.9 (89.1-94.0) | 95.5 (93.3-97.0) | 92.4 (89.7-94.4) | 95.8 (93.6-97.3) |
| 4 | 173 | 5 | 122 | 10 | 96.1 (93.3-97.7) | 97.2 (94.7-98.5) | 92.4 (88.9-94.9) | 94.5 (91.4-96.6) |
| 5 | 116 | 2 | 93 | 7 | 97.9 (95.0-99.1) | 98.3 (95.6-99.4) | 93.0 (88.8-95.7) | 94.3 (90.4-96.7) |
| 6 | 73 | 0 | 74 | 7 | 100.0 (97.6-100.0) | 100.0 (97.6-100.0) | 91.4 (85.9-94.8) | 91.2 (85.7-94.8) |
| 7 | 56 | 0 | 54 | 7 | 100.0 (96.8-100.0) | 100.0 (96.8-100.0) | 88.5 (81.5-93.1) | 88.9 (81.9-93.4) |
| 8 | 37 | 0 | 42 | 5 | 100.0 (95.6-100.0) | 100.0 (95.6-100.0) | 89.4 (81.0-94.3) | 88.1 (79.5-93.4) |

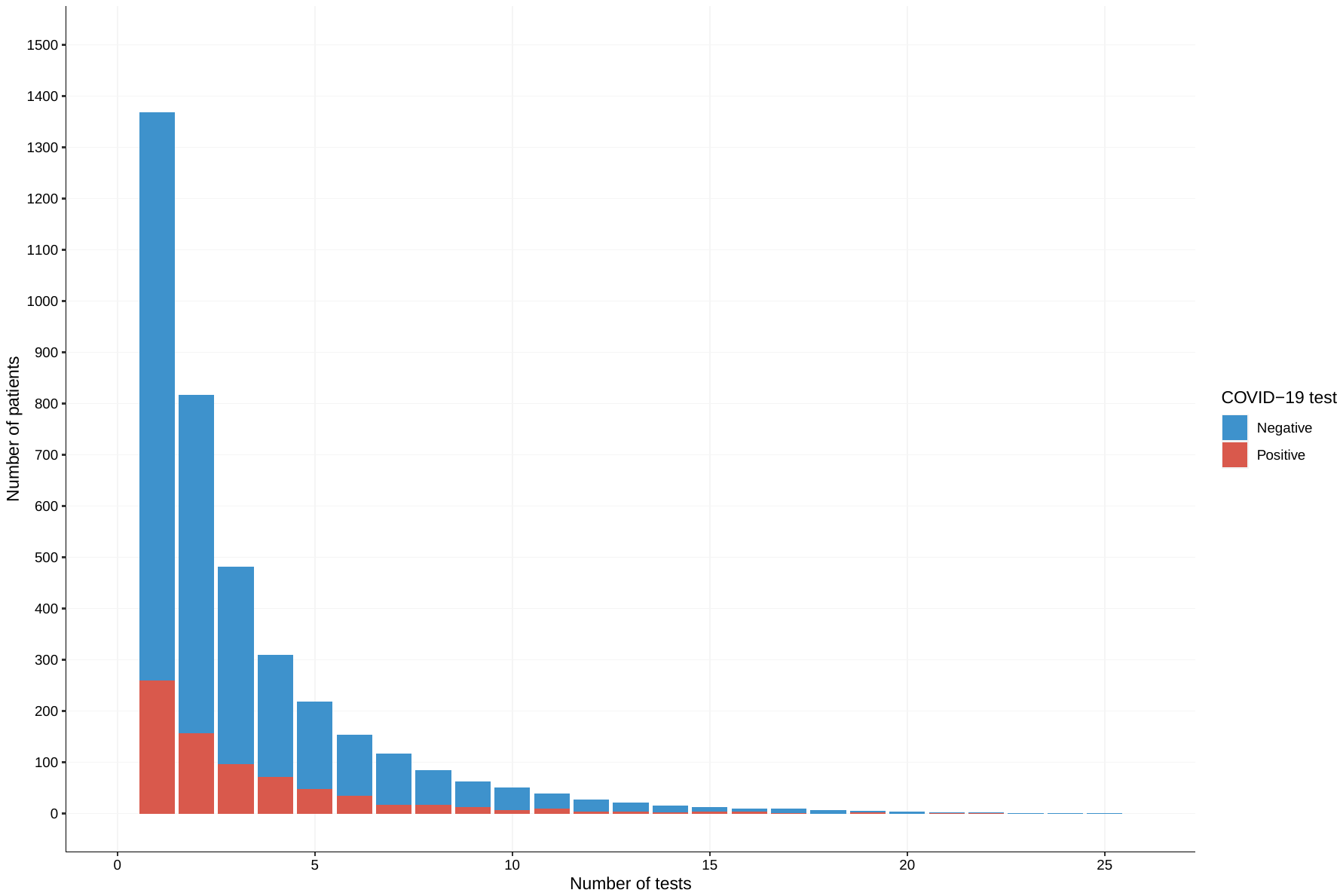
**eFigure 1.** Stack plot of the number of RT-PCR tests performed stratified according to whether the test was positive (red) or negative (blue).

**eFigure 2.** Heat map of RT-PCR testing in patients with confirmed COVID-19 stratified according to whether the index test was negative (a) or positive (b).

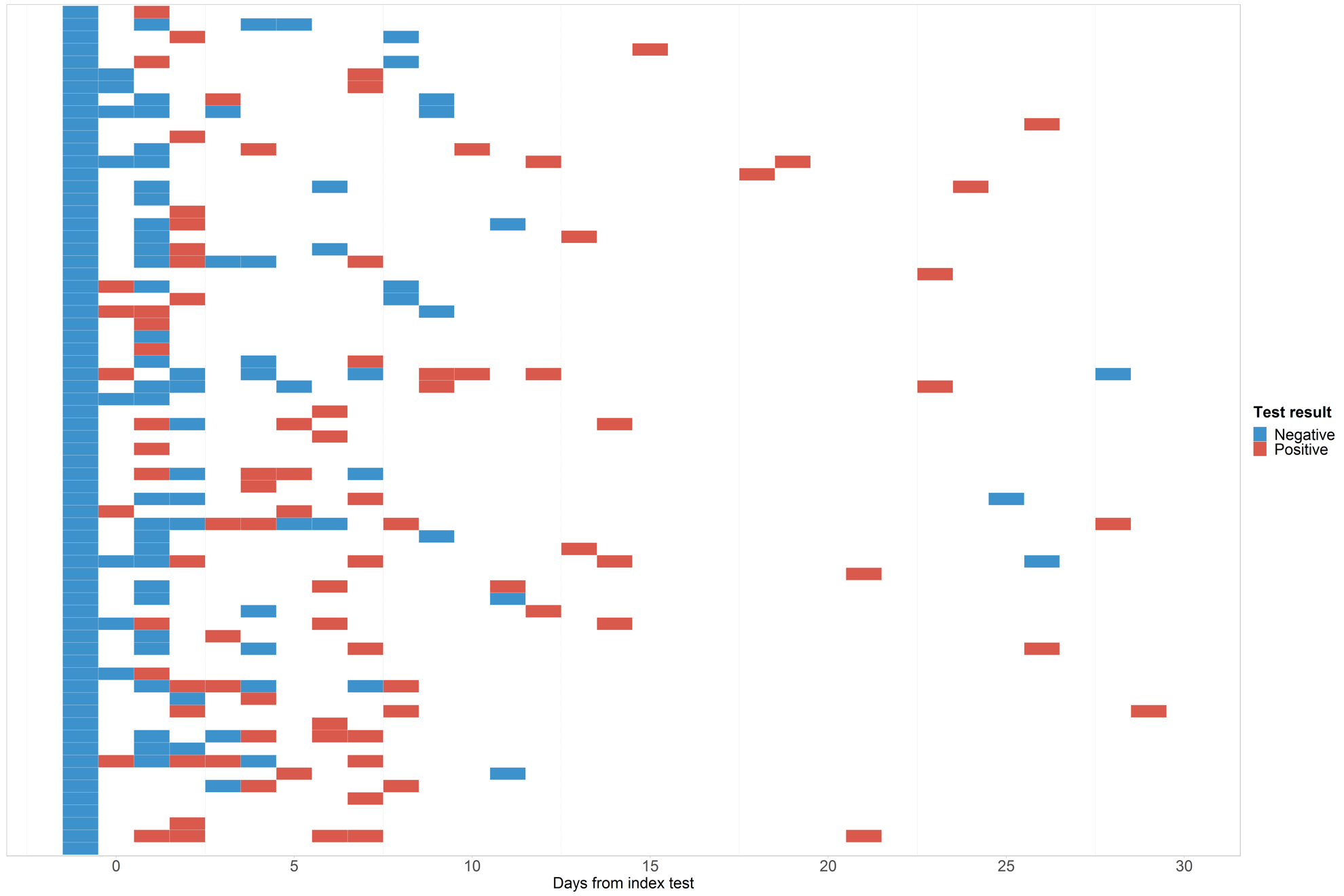
**(a)**

**(b)**

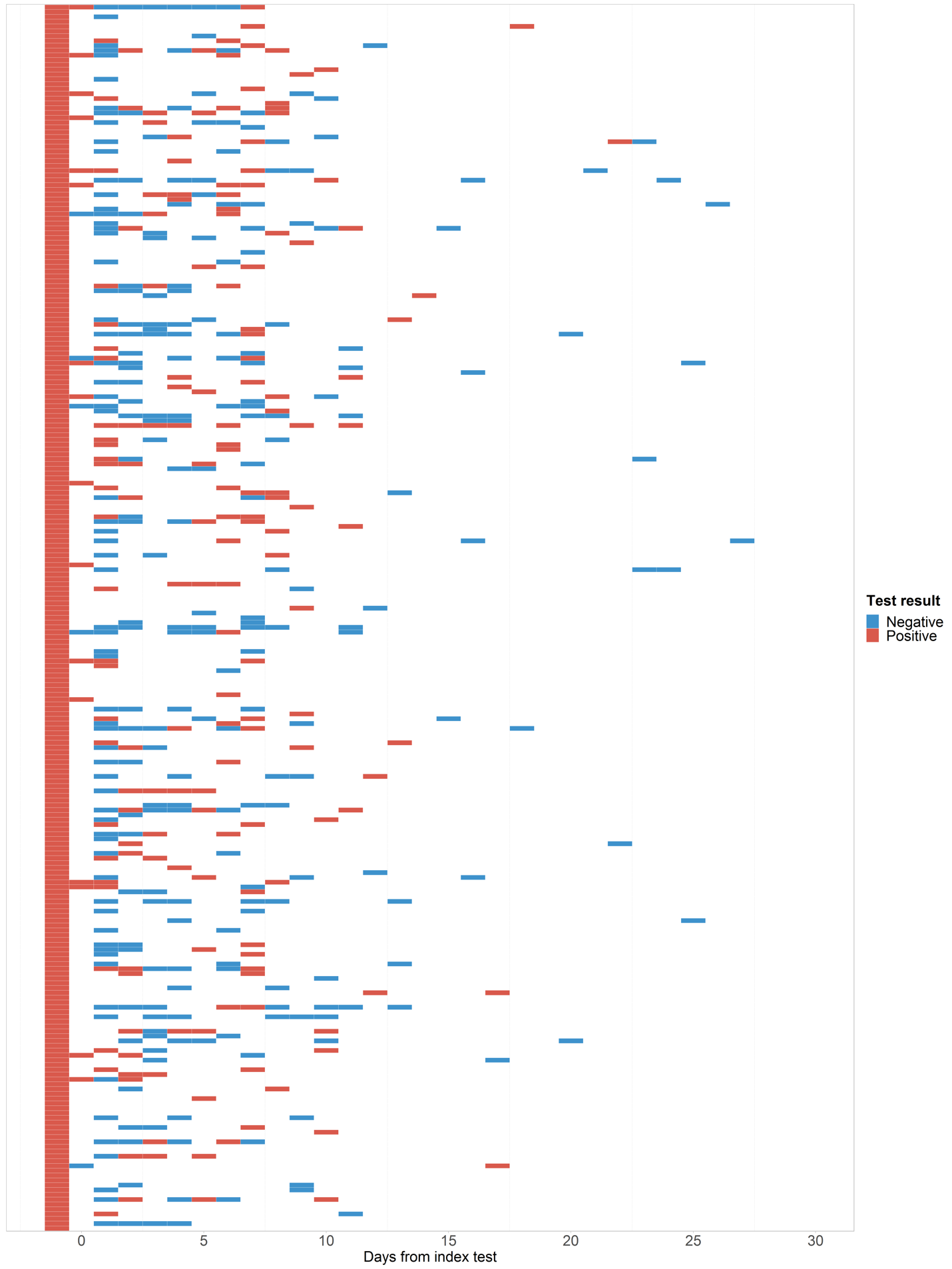

**eFigure 3.** Sensitivity of serial testing using the combined nasal and throat swab for the primary (confirmed and probable COVID-19) and secondary (confirmed COVID-19) outcome in patients who were tested at least four times

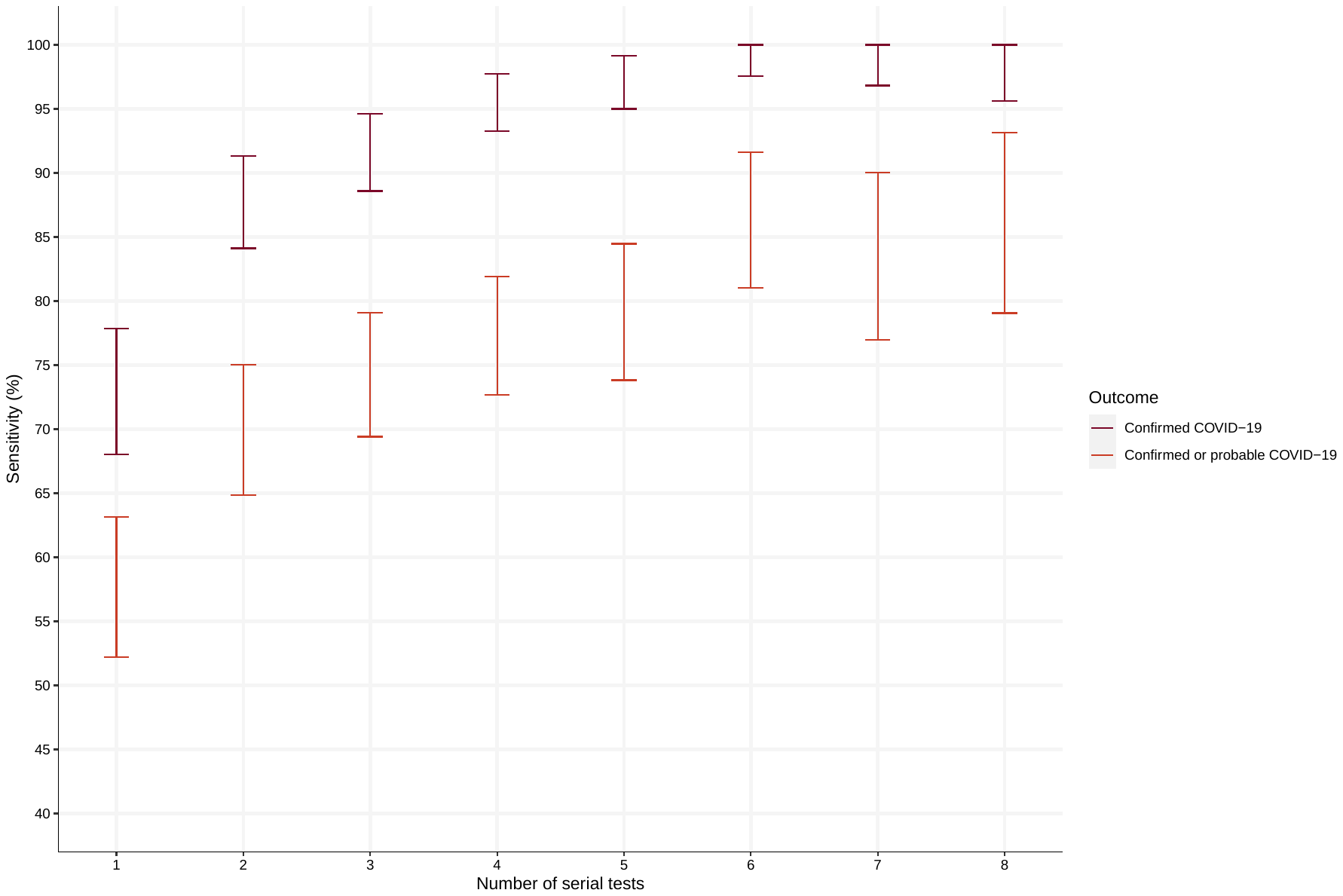

**eFigure 4.** Negative predictive value of serial testing using the combined nasal and throat swab for the primary (confirmed and probable COVID-19) and secondary (confirmed COVID-19) outcome in patients who were tested at least four times

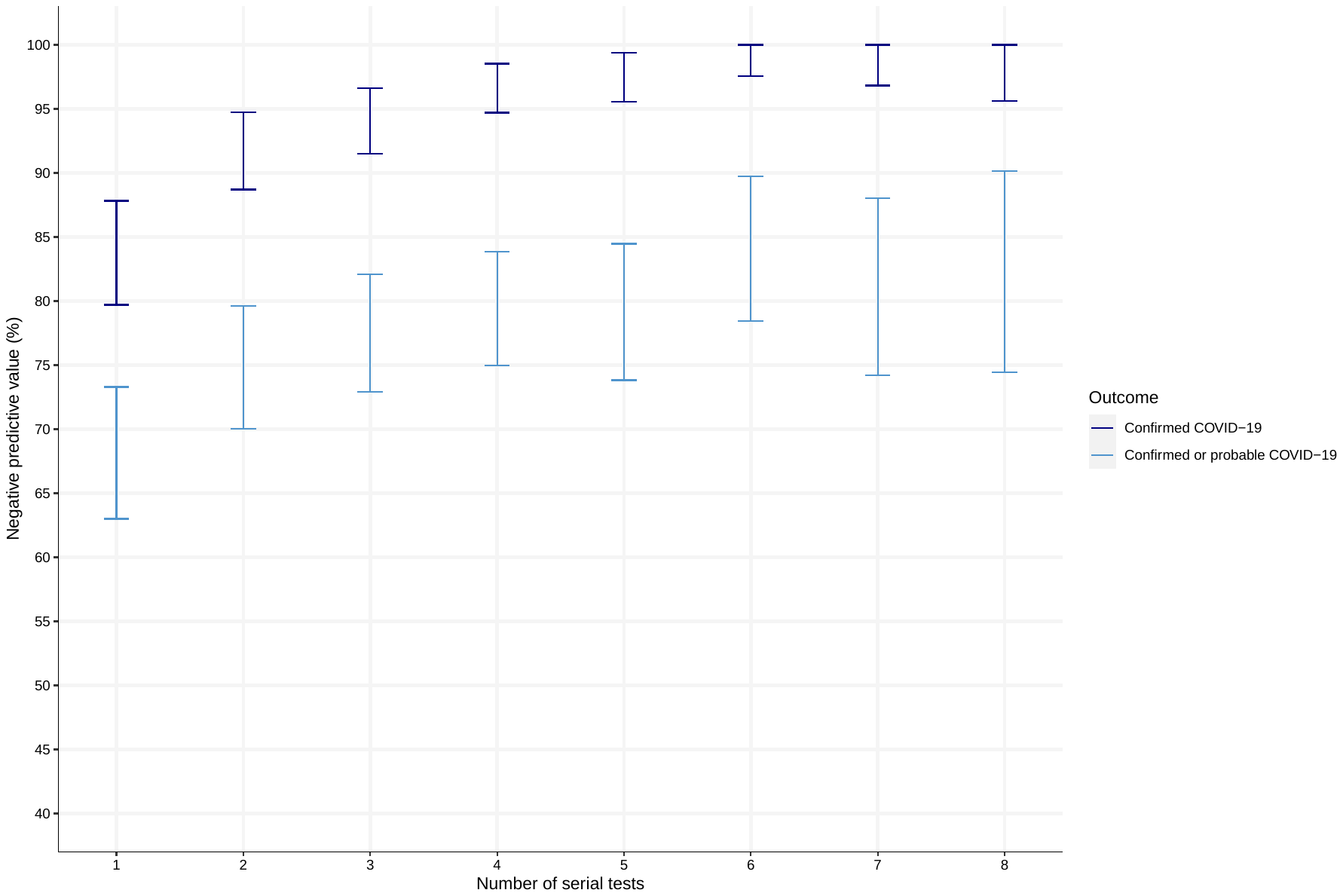

**eFigure 5.** Forest plot of the (a) sensitivity and (b) negative predictive value of the index combined nasal and throat swab for a diagnosis of confirmed COVID-19 stratified by subgroups

## (a)

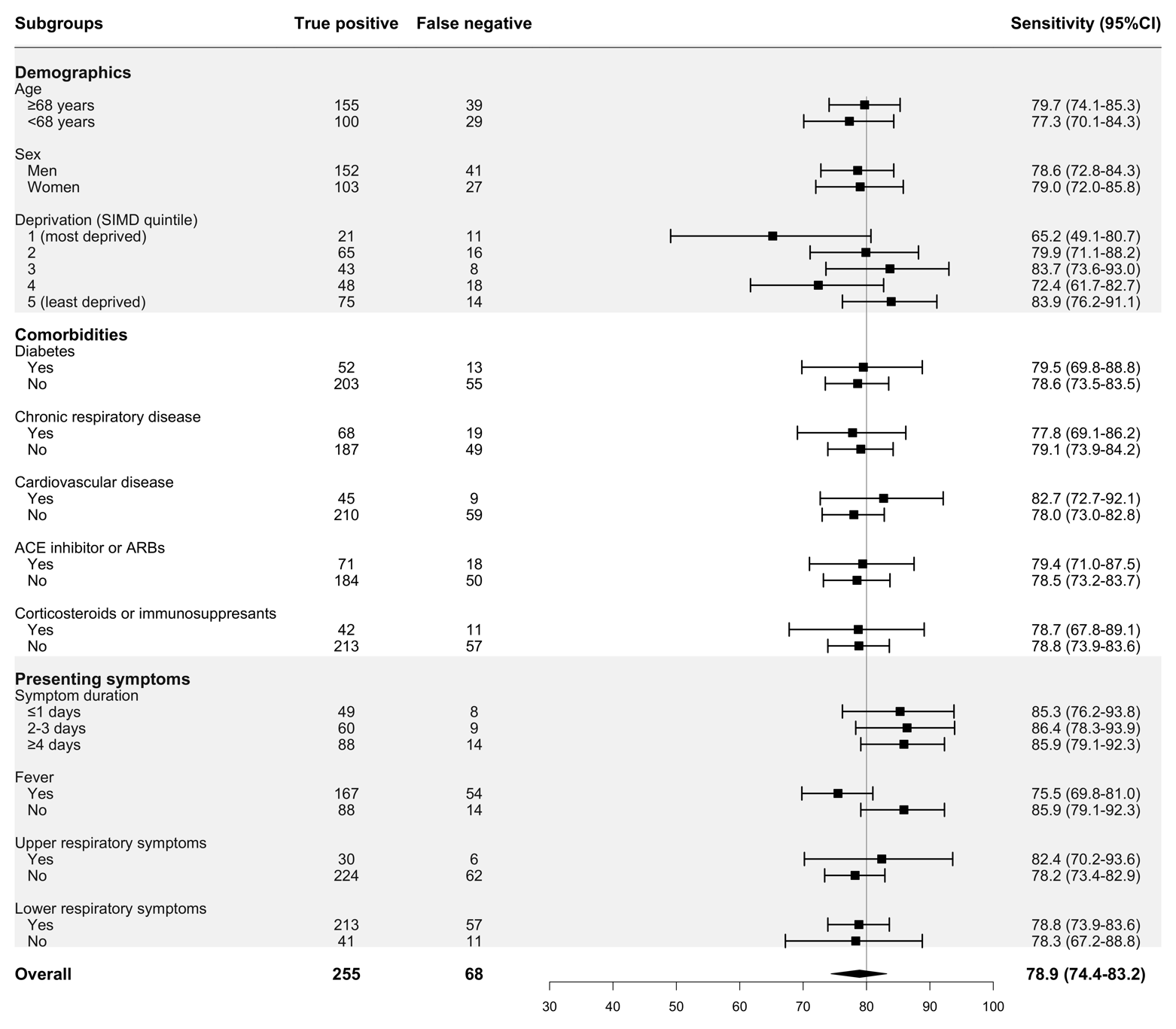

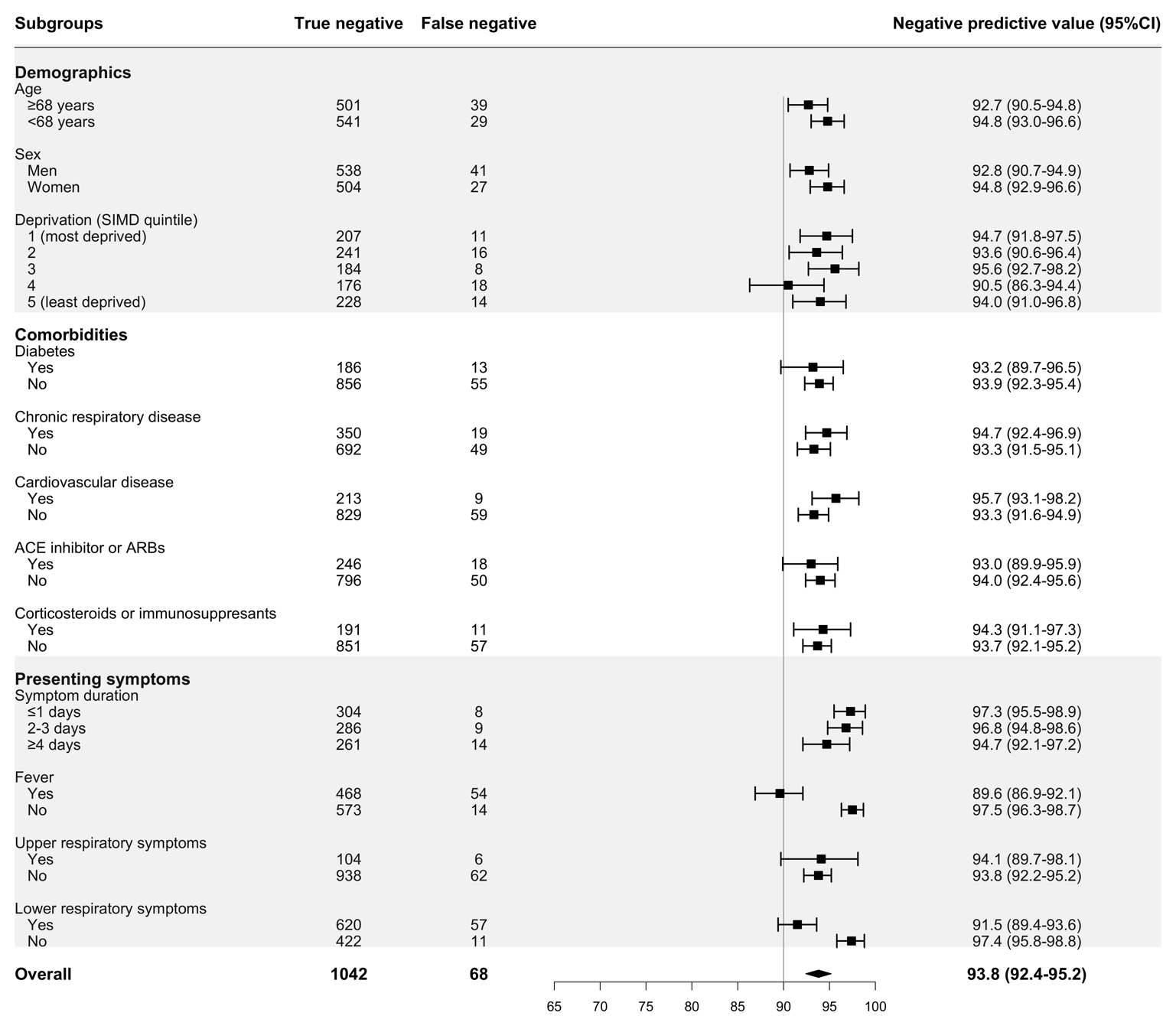
**(b)**
